# Little genomic support for cyclophilin A-matrix metalloproteinase-9 pathway as a therapeutic target for cognitive impairment in *APOE4* carriers

**DOI:** 10.1101/2021.04.22.21255729

**Authors:** Emma L Anderson, Dylan M Williams, Venexia Walker, Neil M Davies

## Abstract

Therapeutic targets for halting the progression of Alzheimers disease pathology are lacking. Recent evidence suggests that APOE4, but not APOE3, activates the cyclophilin A matrix metalloproteinase-9 (CypA MMP9) pathway in the cerebrospinal fluid, leading to an accelerated breakdown of the blood brain barrier (BBB) and thereby causing neuronal and synaptic dysfunction. Furthermore, blockade of the CypA MMP9 pathway in APOE4 knock-in mice restores BBB integrity and subsequently normalizes neuronal and synaptic function. Thus, CypA has been suggested as a potential target for treating APOE4 mediated neurovascular injury and the resulting neuronal dysfunction and degeneration. The odds of drug targets passing through clinical trials are greatly increased if they are supported by genomic evidence. We found little evidence that CypA or MMP9 affected the risk of Alzheimers disease or cognitive impairment using two-sample Mendelian randomization and polygenic risk score analysis in humans. This casts doubt on whether they are likely to represent effective drug targets for cognitive impairment in APOE4 carriers.

---

Therapeutic targets for halting the progression of Alzheimer’s disease pathology are lacking. Montagne et al^1^ recently reported that *APOE4*, but not *APOE3*, activates the cyclophilin A-matrix metalloproteinase-9 (CypA-MMP9) pathway in the cerebrospinal fluid, arguing that this led to an accelerated breakdown of the blood-brain barrier (BBB) and thereby causing neuronal and synaptic dysfunction. The authors have previously shown that blockade of the CypA–MMP9 pathway in *APOE4* knock-in mice restores BBB integrity and subsequently normalizes neuronal and synaptic function^2^. Thus, CypA is suggested as a potential target for treating APOE4-mediated neurovascular injury and the resulting neuronal dysfunction and degeneration. The odds of drug targets passing through clinical trials are greatly increased if they are supported by genomic evidence ^3^. We found little evidence that the drug targets identified by Montagne et al. affected the risk of Alzheimer’s disease using two-sample Mendelian randomization and polygenic risk score analysis in humans. This casts doubt on whether they are likely to represent effective drug targets for cognitive impairment in *APOE4* carriers.

We used two-sample Mendelian randomization^4^ to examine (i) whether *APOE4* (tagged by the C allele of single nucleotide polymorphism rs429358 ^5^) has a causal effect on blood-based CypA or MMP9 expression quantitative trait loci (eQTLs) and protein quantitative trait loci (pQTLs), and (ii) whether blood-based CypA and MMP9 eQTLs and pQTLs have a causal effect on the risk of Alzheimer’s disease (AD). We used the largest publicly available blood-based eQTL genome-wide association study (GWAS) meta-analysis (eQTLGen; n=31,684^6^), pQTL GWAS meta-analysis (n=3,301^7^) and Alzheimer’s disease GWAS meta-analysis (n=71,880 cases and 383,378 controls^8^). Full details of the harmonization procedure and statistical methods can be found in the supplement.

Secondly, we examined whether *APOE4* (C allele of rs429358) and polygenic risk scores for CypA and MMP9 had a causal effect on (i) Alzheimer’s disease-by-proxy (parental dementia) and (ii) continuous markers of cognitive function in the UK Biobank: visual memory (n= 336,679), reaction time (n=335,019) and fluid intelligence score (n=120,109). We examined these effects both in the total sample and in age-stratified tertiles to examine potential age-dependent effects. We also examined whether any of the observed effects were modified by *APOE4* status (i.e. do CypA and MMP9 eQTLs and pQTLs affect markers of cognitive function differently in *APOE4* carriers vs non-carriers). Full details of the statistical analysis are in the supplement.

We found little evidence to suggest *APOE4* has a causal effect on CypA or MMP9 eQTLs or pQTLs using two-sample Mendelian randomization. Confidence intervals were wide and could not exclude a meaningful effect in either direction (Table 2 of the supplement). There was little evidence that CypA or MMP9 eQTLs or pQTLs affected the odds of Alzheimer’s disease, and these estimates were very precise (Table 3 of the supplement). In individual-level analyses in the UK Biobank, *APOE4* carrier status was strongly associated with odds of reporting one or both parents to have dementia (Table 4 of the supplement) and there was evidence of an age-dependent effect on fluid intelligence scores (scores worsening with age, Figure 1). *APOE4* carrier status had no notable causal effect on visual memory scores or reaction times (Figure 1). Overall, there was no consistent evidence to suggest a causal effect of CypA or MMP9 eQTL or pQTL PRSs on odds of reporting one or both parents to have dementia (Table 4 of supplement) or on any of the continuous markers of cognitive function (Figure 1). Higher CypA eQTL and pQTL PRSs were associated consistently with faster reaction times in only the middle age tertile. A higher MMP9 eQTL PRS was associated with lower fluid intelligence scores in the youngest tertile only, and with slower reaction times in the middle and oldest age tertiles. Conversely, a higher MMP9 pQTL PRS was not associated with fluid intelligence scores in any age tertile and was associated with faster reaction times in only the middle tertile. Finally, there was little consistent evidence to suggest that causal effects of CypA and MMP9 eQTLs and pQTLs on the continuous cognitive function markers were modified by APOE4 carrier status (Figure 2). In the oldest age tertile, greater MMP9 eQTL polygenic risk scores were unexpectedly associated with better fluid intelligence scores and reaction times in homozygous *APOE4* carriers, and greater CypA pQTL polygenic risk scores were associated with better fluid intelligence scores, but worse visual memory in homozygous *APOE4* carriers. It is worth noting that these findings were comparable in the whole sample (rather than restricting to the oldest age tertile, results not shown).

**Figure 1:**
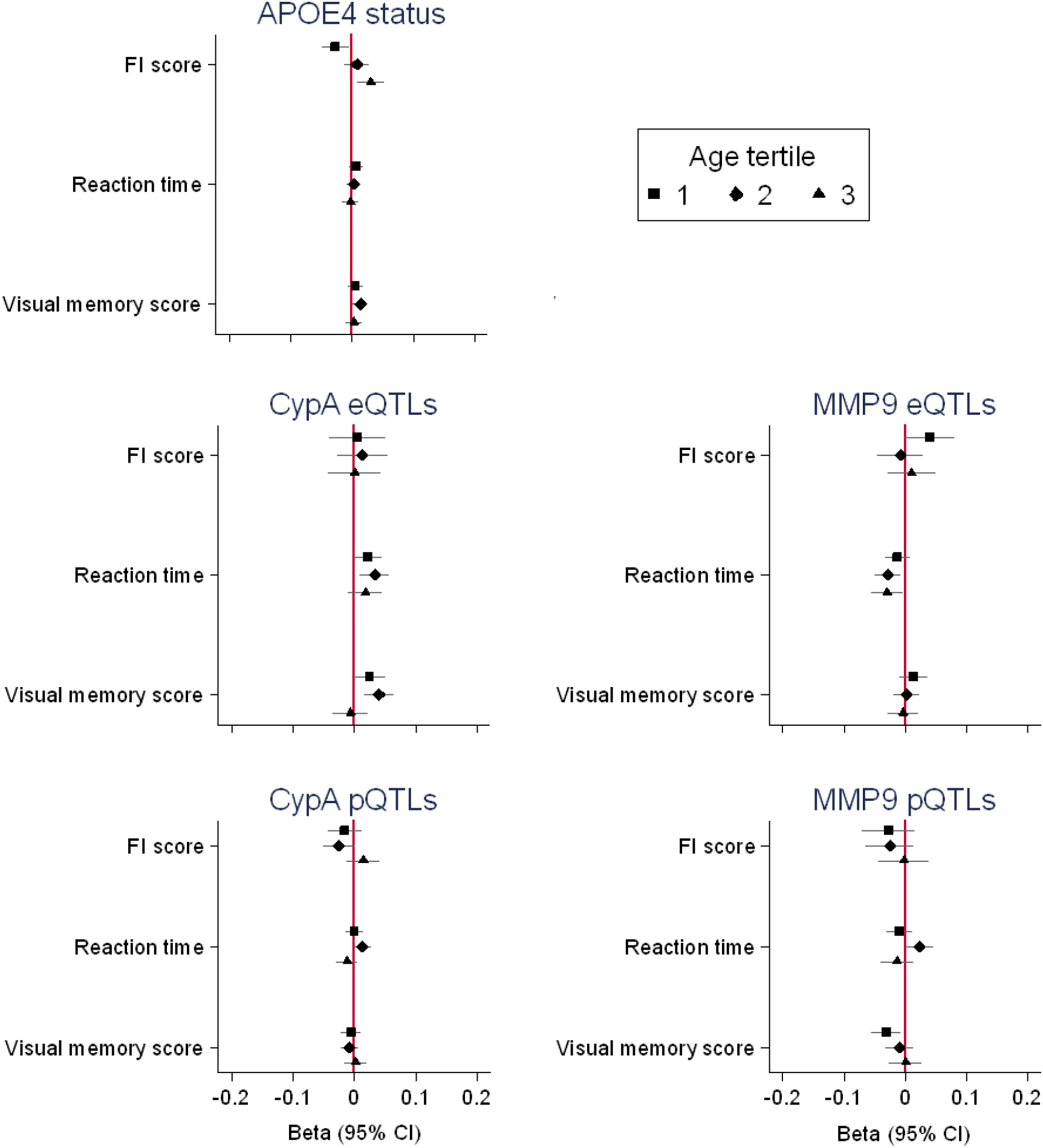
Causal effects of APOE4 carrier status (carrier vs non-carrier) and CypA and MMP9 eQTL and pQTL PRSs on the continuous cognitive function outcomes in the UK Biobank, by age tertiles. All outcomes are in standard deviation units and higher values represent poorer performance.

**Figure 2:**
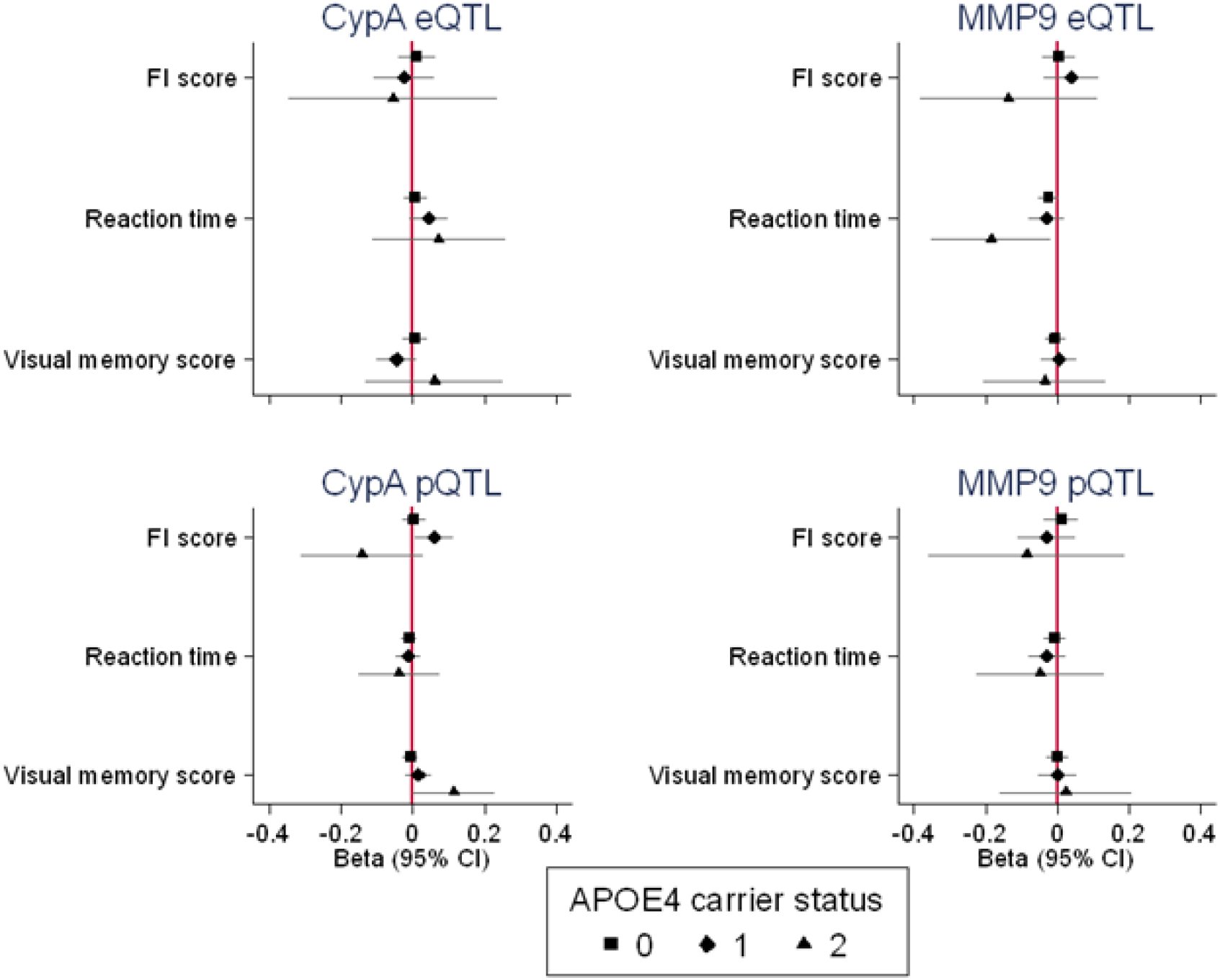
Causal effects of CypA and MMP9 eQTL and pQTL PRSs on the continuous cognitive function outcomes in the UK Biobank (fluid intelligence score, reaction time and visual memory) in the oldest tertile, by APOE4 carrier status. All outcomes are in standard deviation units and higher values represent poorer performance.

In conclusion, we found very little genomic support for the suggestion by Montange et al that CypA is a promising therapeutic target for cognitive impairment in *APOE4* carriers. Causal effect estimates from the Mendelian randomization analyses were very precisely null, and there was no consistent evidence from the individual analyses on continuous cognitive function markers in the UK Biobank. Although statistical power is lower in the UK Biobank analysis compared to the Mendelian randomization analyses, any plausible effects are likely to be extremely small and not clinically meaningful given the confidence intervals estimated. The analyses presented here were conducted on the largest available samples in humans, and we were able to examine a series of related outcomes. There are, of course, limitations to these results. Firstly, CypA and MMP9 eQTLs and pQTLs are from blood and not specifically brain tissue. However, recent evidence^9^ shows brain eQTLs from different brain regions to have very low correlations; indeed some brain regions correlate more strongly with blood and kidney eQTLs than they do with other brain regions. Thus, using eQTLs from brain tissue is not necessarily a useful way to overcome that limitation (as it largely depends on the region analysed) and they are also underpowered (as fewer brain samples exist compared to other tissue samples). It is also worth noting that drugs used to inhibit CypA accumulate in brain microvessels, but do not cross the blood-brain barrier, suggesting blood is likely a relevant tissue2. Secondly, the UK Biobank population is relatively young with respect to the Alzheimer’s disease course, and we may have been able to detect causal effects in a much older population. That said, there was no evidence of causal effects in the Mendelian randomization analyses which used the largest case-control Alzheimer’s disease GWAS as the outcome, and individual-level results from the UK Biobank looked very similar when restricted to the oldest tertile (age 63-72 years), compared to the youngest and middle age tertile or the whole sample. Finally, It is worth noting that pleiotropy is an unlikely explanation for these findings as it would tend to bias away from the null. These results are a useful additional source of evidence about the relationship between CypA and MMP9 and Alzheimer’s disease risk, which could help prioritise effective therapeutic targets to help reduce the burden of Alzheimer’s disease.

## Data Availability

eQTL and pQTL genome-wide association study summary statistics are available on the MR-Base platform. Alzheimer's disease summary statistics are available from the following website: https://ctg.cncr.nl/software/summary_statistics

## References

1 Montagne, A. et al. APOE4 leads to blood-brain barrier dysfunction predicting cognitive decline. Nature 581, 71–76, doi:10.1038/s41586-020-2247-3 (2020).

2 Bell, R. D. et al. Apolipoprotein E controls cerebrovascular integrity via cyclophilin A. Nature 485, 512–516, doi:10.1038/nature11087 (2012).

3 King, E. A., Davis, J. W. & Degner, J. F. Are drug targets with genetic support twice as likely to be approved? Revised estimates of the impact of genetic support for drug mechanisms on the probability of drug approval. PLoS Genet 15, e1008489, doi:10.1371/journal.pgen.1008489 (2019).

4 Davey Smith, G. & Hemani, G. Mendelian randomization: genetic anchors for causal inference in epidemiological studies. Hum Mol Genet 23, R89–98, doi:10.1093/hmg/ddu328 (2014).

5 Babenko, V. N. et al. Haplotype analysis of APOE intragenic SNPs. BMC Neurosci 19, 16, doi:10.1186/s12868-018-0413-4 (2018).

6 Võsa, U. et al. Unraveling the polygenic architecture of complex traits using blood eQTL metaanalysis. bioRxiv, 447367, doi:10.1101/447367 (2018).

7 Zheng, J. et al. Phenome-wide Mendelian randomization mapping the influence of the plasma proteome on complex diseases. Nat Genet 52, 1122–1131, doi:10.1038/s41588-020-0682-6 (2020).

8 Jansen, I. E. et al. Genome-wide meta-analysis identifies new loci and functional pathways influencing Alzheimer’s disease risk. Nat Genet 51, 404–413, doi:10.1038/s41588-018-0311-9 (2019).

9 de Klein, N. et al. Brain expression quantitative trait locus and network analysis reveals downstream effects and putative drivers for brain-related diseases. bioRxiv, 2021.2003.2001.433439, doi:10.1101/2021.03.01.433439 (2021).

