## Supplemental Material for "Little genomic support for cyclophilin A-matrix metalloproteinase-9 pathway as a therapeutic target for cognitive impairment in *APOE4* carriers"

Mendelian Randomization analysis methods………………………………………………………………….2

UK Biobank analyses methods.……………………………………………………………………………………….4

Mendelian Randomization results…………………………………………………………………………………..7

UK Biobank results – parental dementia………………………………………………………………………….7

**Mendelian Randomization analysis methods**
Methods for conducting two-sample MR analyses have been published previously^4^. Briefly, two-sample MR provides an estimate of the causal effect of an exposure on an outcome, using independent samples to obtain the gene-exposure and gene-outcome associations, provided three key assumptions hold: (i) genetic variants are robustly associated with the exposure of interest (i.e. replicate in independent samples), (ii) there is no confounding of the causal effect of the genetic variants on the outcome (for example, by population stratification (ref)) and (iii) there are no effects of the genetic variants on the outcome, independent of the exposure (i.e. no horizontal pleiotropy)(ref).

**Data**
Expression quantitative trait loci (eQTLs)^6^ and protein quantitative trait loci (pQTLs)^7^ genome wide association studies (GWAS) have been previously conducted. For the outcome, we used the largest available GWAS meta-analysis of AD (n= 71,880 clinically diagnosed AD and AD-by-proxy cases, 383,378 controls)^8^. All GWASs used in these analyses were conducted primarily on participants of European ancestry. Ethics approval was obtained by the original studies. F statistics are provided in the results tables. F statistics provide an indication of instrument strength and are a function of how much variance in the trait is explained by the set of genetic instruments being used, the number of genetic instruments being used, and the sample size. F statistics greater than 10 indicate that the analysis is unlikely to suffer from weak instrument bias^10^. All F statistics were above 10.
**Harmonization procedure and statistical analysis**
MR-Base ([www.mrbase.org](http://www.mrbase.org))^11^ was employed to perform all MR analyses. To estimate causal effects of APOE4 on CypA and MMP9 eQTLs and pQTLs, the E4 allele of APOE was tagged by the presence of cysteine at rs429358 (one C for heterozygotes, two for homozygotes). SNP (rs429358)-outcome estimates were extracted from the CypA and MMP9 eQTL and pQTL GWASs and effects of APOE4 on the outcomes are shown in Table 2 below. For causal effects of CypA and MMP9 eQTLs and pQTLs on Alzheimer’s disease, one approximately independent genome-wide significant (p<5x10^-8^) single nucleotide polymorphism (SNP) was identified as being associated with CypA eQTLs; eight SNPs were associated with MMP9 eQTLs; one SNP with CypA pQTLs and three SNPs with MMP9 pQTLs, in two recent meta-analyses described above. Details of these SNPs are provided in Table 1 below.  SNP-outcome estimates for all SNPs were extracted from the Alzheimer’s disease GWAS described above. No SNPs were excluded due to low minor allele frequencies (<1%) and all studies included in the analyses were coded on the forward strand, thus, no palindromic SNPs were excluded from analyses*.* In an MR analysis, the effect of a SNP on exposure and an outcome must be harmonised to be relative to the same allele. SNPs for the exposure were coded so that the effect allele was always the ‘increasing allele’ (i.e. increasing CypA and MMP9 eQTLs), and the alleles were harmonized so that the effect on the outcome corresponded to the same allele as the exposure.

All SNP-exposure estimates are in standard deviation (SD) units. SNP-outcome estimates are in units of log odds ratios (ORs) for AD. Coefficients were combined using an inverse-variance-weighted (IVW) approach to give an overall estimate of the causal effect across all SNPs included for each eQTL and pQTL. The estimator is a Wald ratio and is equivalent to a weighted regression of the SNP-outcome coefficients on the SNP-exposure coefficients with the intercept constrained to zero. For causal effects of CypA and MMP9 eQTLs and pQTLs on Alzheimer’s disease, results were exponentiated to ORs, thus, causal effect estimates are interpreted as the odds of AD per standard deviation increase in CypA and MMP9 eQTLs or pQTLs.

**UK Biobank analysis methods
Data**
We conducted a polygenic risk score analysis in the UK Biobank examining causal effects of CypA and MMP9 on by-proxy AD, visual memory, reaction time and fluid intelligence. The UK Biobank is a large-scale population-based study that recruited 502,616 individuals, aged 40-69 years old, between 2006-2010. The study was established to enable investigations of genetic and nongenetic determinants of diseases of middle and old age ^6 7^. The UK Biobank was approved by the North West Research Ethics committee. Genotyping was completed for all participants at baseline. Participants also reported whether their mother and/or father had dementia. At the initial visit, reaction time, visual memory and fluid intelligence scores were assessed. Reaction time was assessed using 12 rounds of the card-game 'Snap’. The participant was shown two cards at a time; if both cards were the same, they pressed a button-box that was on the table in front of them as quickly as possible.  Thus, higher values equal poorer performance. Visual memory was assessed using incorrect matches on a pairs matching test. Participants were asked to memorise the position of as many matching pairs of cards as possible. The cards were then turned face down on the screen and the participant was asked to touch as many pairs as possible in the fewest tries. Incorrect matches were counted, thus higher values equal poorer performance. Fluid intelligence was examined in a subsample of participants, by answering a series of questions designed to assess their capacity to solve problems that require logic and reasoning ability, independently of acquired knowledge. The participant had 2 minutes to complete as many questions as possible from the test, and higher scores represent better performance.

**Statistical analysis**

The current study sample was restricted to those of “White British” origin, who were unrelated, did not report being adopted, and with no missing data for all covariables. Weighted polygenic risk scores were generated for CypA and MMP9 eQTLs and pQTLs (i.e. four different polygenic risk scores), for each participant with genetic data. They included all SNPs associated with CypA and MMP9 eQTLs and pQTLs at genome-wide significance (p≤5×10^-8^), clumped at R^2^=0.001 and a 10,000kb window (GWAS refs). Polygenic risk scores were calculated using PLINK (version 2.0). Each score was calculated from the effect size-weighted sum of associated alleles within each participant.

Fluid intelligence scores were approximately normally distributed. Reaction times and visual memory scores were natural log transformed to approximate a normal distribution. A constant of +1 was added to all visual memory scores to ensure no zeros were log transformed. Parental AD was coded as 0, 1 or 2 parents with AD. Thus, casual effects of the polygenic risk scores on all continuous outcomes were estimated using linear regression models and ordinal logistic regression models were used when parental AD was the outcome. All regression models were adjusted for age and sex. As higher values for reaction time and visual memory (incorrect pair matches) represent poorer performance, we reversed fluid intelligence scores so that they were in a consistent direction with the other continuous outcomes (i.e. higher fluid intelligence scores represent poorer performance). We first conducted these analyses on the whole UK Biobank sample. We then examined these effects within age-stratified tertiles to interrogate potential age-dependent effects. The age-stratified analysis was conducted because, on average, the UK Biobank population is relatively young with respect to AD diagnoses, and if a proportion of the sample have underlying AD pathology, we would anticipate this proportion to be greatest in the oldest tertile. Thus, it is plausible that we may only observe effects of the polygenic risk scores on cognitive impairment indicators in the population which have underlying AD pathology (i.e. the middle to oldest age tertiles). Finally, we examined whether any effects of the polygenic risk score on cognitive impairment indicators were modified by APOE4 carrier status (zero, one or two C alleles for SNP rs429358).

**Table 1: Instruments for CypA and MMP9 eQTLs and pQTLs**

| **Trait** | **RSID** | **Effect allele** | **Other Allele** | **Effect allele frequency** | **Beta** | **Standard error** | **P** |
| --- | --- | --- | --- | --- | --- | --- | --- |
| **CypA eQTL** | rs6463247 | T | C | 0.816911 | 0.417215 | 0.0149835 | 1.2388e-170 |
| **MMP9 eQTLs** | rs3731827 | C | T | 0.418566 | 0.096111 | 0.012037 | 1.41E-15 |
|  | rs7613595 | C | A | 0.23545 | 0.109595 | 0.013998 | 4.89E-15 |
|  | rs149110519 | T | C | 0.052616 | -0.16455 | 0.026622 | 6.37E-10 |
|  | rs56388170 | T | G | 0.289189 | -0.14 | 0.013074 | 9.38E-27 |
|  | rs149007767 | T | C | 0.149708 | -0.14688 | 0.016636 | 1.06E-18 |
|  | rs4065321 | T | C | 0.539612 | 0.077578 | 0.011923 | 7.69E-11 |
|  | rs13925 | A | G | 0.148395 | 0.353837 | 0.016475 | 2.56E-102 |
|  | rs6073969 | T | C | 0.934435 | 0.203758 | 0.023984 | 1.97E-17 |
| **CypA pQTL** | rs62143198 | A | G | 0.21453 | 0.6335 | 0.0287 | 2.86E-108 |
| **MMP9 pQTLs** | rs398076 | A | G | 0.41095 | 0.156 | 0.0255 | 4.75E-10 |
|  | rs62143194 | C | G | 0.77636 | -0.1974 | 0.0306 | 5.56E-11 |
|  | rs2250889 | C | G | 0.95338 | -0.6221 | 0.0585 | 1.03E-26 |

**Results**

**Mendelian randomization analyses**

**Table 2: Association of APOE (rs429358) with CypA and MMP9 eQTLs and pQTLs**

|  | **Beta (95% CI)** | **P** |
| --- | --- | --- |
| **CypA eQTLs** | -0.01 (-0.05 to 0.03) | 0.62 |
| **MMP9 eQTLs** | 0.001 (-0.04 to 0.04) | 0.97 |
| **CypA pQTLs** | 0.02 (-0.04 to 0.08) | 0.55 |
| **MMP9 pQTLs** | 0.003 (-0.06 to 0.06) | 0.93 |

Causal effect estimates are interpreted as the average difference in CypA and MMP9 eQTLs and pQTLs per risk increasing allele

**Table 3: Causal effects of CypA and MMP9 eQTLs and pQTLs on risk of Alzheimer’s disease**

|  | **N SNPs** | **F statistics** | **OR (95% CI)** | **P** |
| --- | --- | --- | --- | --- |
| **CypA eQTLs** | 1 | 775.34 | 1.00 (0.99 to 1.01) | 0.80 |
| **MMP9 eQTLs** | 8 | 116.45 | 1.00 (0.98 to 1.01) | 0.45 |
| **CypA pQTLs** | 1 | 487.22 | 1.00 (0.99 to 1.01) | 0.82 |
| **MMP9 pQTLs** | 3 | 64.04 | 1.00 (0.98 to 1.01) | 0.51 |

Causal effect estimates are interpreted as the odds of Alzheimer’s disease per standard deviation increase in CypA and MMP9 eQTLs or pQTLs.

**UK Biobank results**

**Table 4: Causal effects of APOE4 and polygenic risk scores for CypA and MMP9 eQTLs and pQTLs with odds of reporting one or both parents to have dementia (n=328,509)**

|  | **AD-by-proxy** | |
| --- | --- | --- |
|  | **OR (95% CI)** | **P** |
| **APOE4** | 25.77 (23.08 to 28.77) | <0.0001 |
| **CypA eQTLs** | 1.00 (0.96 to 1.05) | 0.89 |
| **MMP9 eQTLs** | 1.03 (0.99 to 1.07) | 0.22 |
| **CypA pQTLs** | 1.01 (0.98 to 1.04) | 0.51 |
| **MMP9 pQTLs** | 0.99 (0.95 to 1.03) | 0.64 |

Causal effect estimates are interpreted as the odds of reporting one or both parents as having dementia per risk increasing allele of APOE4, and per standard deviation increase in CypA and MMP9 eQTLs or pQTLs.

1	Montagne, A. *et al.* APOE4 leads to blood-brain barrier dysfunction predicting cognitive decline. *Nature* **581**, 71-76, doi:10.1038/s41586-020-2247-3 (2020).

2	Bell, R. D. *et al.* Apolipoprotein E controls cerebrovascular integrity via cyclophilin A. *Nature* **485**, 512-516, doi:10.1038/nature11087 (2012).

3	King, E. A., Davis, J. W. & Degner, J. F. Are drug targets with genetic support twice as likely to be approved? Revised estimates of the impact of genetic support for drug mechanisms on the probability of drug approval. *PLoS Genet* **15**, e1008489, doi:10.1371/journal.pgen.1008489 (2019).

4	Davey Smith, G. & Hemani, G. Mendelian randomization: genetic anchors for causal inference in epidemiological studies. *Hum Mol Genet* **23**, R89-98, doi:10.1093/hmg/ddu328 (2014).

5	Babenko, V. N. *et al.* Haplotype analysis of APOE intragenic SNPs. *BMC Neurosci* **19**, 16, doi:10.1186/s12868-018-0413-4 (2018).

6	Võsa, U. *et al.* Unraveling the polygenic architecture of complex traits using blood eQTL metaanalysis. *bioRxiv*, 447367, doi:10.1101/447367 (2018).

7	Zheng, J. *et al.* Phenome-wide Mendelian randomization mapping the influence of the plasma proteome on complex diseases. *Nat Genet* **52**, 1122-1131, doi:10.1038/s41588-020-0682-6 (2020).

8	Jansen, I. E. *et al.* Genome-wide meta-analysis identifies new loci and functional pathways influencing Alzheimer's disease risk. *Nat Genet* **51**, 404-413, doi:10.1038/s41588-018-0311-9 (2019).

9	de Klein, N. *et al.* Brain expression quantitative trait locus and network analysis reveals downstream effects and putative drivers for brain-related diseases. *bioRxiv*, 2021.2003.2001.433439, doi:10.1101/2021.03.01.433439 (2021).

10	Burgess, S., Thompson, S. G. & Collaboration, C. C. G. Avoiding bias from weak instruments in Mendelian randomization studies. *Int J Epidemiol* **40**, 755-764, doi:10.1093/ije/dyr036 (2011).

11	Hemani G. *et al*. MR-Base: a platform for systematic causal inference across the phenome using billions of genetic associations. *bioRxiv* (2017).
